# Managing AI-Enabled Uncertainty in Clinical AI Deployment: Mixed-Methods Study of Governance, Workflow, and Organizational Learning in an ICU Decision Support Pilot

**DOI:** 10.64898/2026.02.04.26345355

**Authors:** Alexander Althammer, Anton Hummel, Jan-Philipp Steghöfer, Felix Reichel, Jonas Kolonko, Sarah Hatfield, Maximilian Fischer, Kerstin Schlögl-Flierl, Paula Ziethmann, Manfred Weiß, Philipp Simon, Markus Mögerlein, Eugen Mamtschur, Oliver Spring, Sergey Shmygalev, Natalia Ortmann, Johannes Raffler, Christian Hinske, Jens O. Brunner, Axel R. Heller, Christina C. Bartenschlager

**Affiliations:** Anaesthesiology and Intensive Care Medicine, University of Augsburg, Augsburg, Germany; XITASO GmbH IT & Software Solutions, Augsburg, Germany; School of Business, Augsburg Technical University of Applied Sciences, Augsburg, Germany; Center for Responsible AI Technologies, Augsburg, Germany; Institute for Employment Research (IAB), Nuremberg, Germany; Department of Medical Informatics, University Hospital of Augsburg, Augsburg, Germany; Digital Medicine, University Hospital of Augsburg, Augsburg, Germany; Faculty III - Economics, Business Informatics and Business Law, University of Siegen, Germany; Department of Technology, Management, and Economics, Technical University of Denmark, Denmark; Health Care Operations/Health Information Management, Faculty of Business and Economics, Faculty of Medicine, University of Augsburg, Augsburg, Germany; Applied Data Science in Health Care, Nürnberg School of Health, Ohm University of Applied Sciences, Nuremberg, Germany

**Keywords:** Artificial intelligence, digital health management, clinical decision support systems, AI governance, learning health systems, implementation evaluation, human-AI interaction, intensive care, workflow, strategic decision-making

## Abstract

**Background:** Health care organizations are increasingly required to make strategic decisions about artificial intelligence (AI) systems before their clinical value, operational consequences, governance requirements, and workforce implications are fully known. Clinical AI pilots can reduce this uncertainty only if they generate management-relevant evidence beyond retrospective model performance, including evidence on workflow fit, governance architecture, user interaction, accountability, and continuous learning.

**Objective:** This study aimed to examine how a prospective intensive care unit (ICU) deployment of an AI-based clinical decision support system (CDSS) surfaced organizational and governance-related uncertainties, and to derive a transferable management toolbox for early-stage clinical AI deployment.

**Methods:** We conducted a prospective, single-center mixed-methods implementation evaluation of a machine-learning-based CDSS for ICU length-of-stay prediction in a surgical ICU at a German university hospital. The study included 267 consecutive ICU stays and was approved by the LMU Munich Institutional Review Board (Project No. 24-0336) and registered with the German Clinical Trials Register (DRKS00037851). The evaluation combined five management-relevant domains: workflow integration and operational adoption, governance architecture and data-protection effects, live bedside benchmarking as a continuous learning mechanism, embedded ethics and accountability design, and human-AI interaction with user heterogeneity assessed using the Psychological Assessment of AI-based Decision Support Systems instrument.

**Results:** The deployment showed that AI-enabled uncertainty was generated primarily at the interface between the CDSS and its organizational setting. A low-burden “glance-and-judge” workflow enabled routine use among consultants and resident physicians, with usage proportions of 72% (191/267) and 61% (162/267), respectively. Governance requirements led to an air-gapped architecture with once-daily data refreshes, enabling compliant use but creating operational latency; outdated case-list entries occurred in 4 of 148 benchmarked stays (2.7%). Live benchmarking functioned as a feedback loop and supported exploratory model refinement, with CDSS mean absolute error decreasing from 5.95 to 4.12 days after the first model update. Embedded ethics informed onboarding, accountability framing, interface wording, and explainability design. Human-AI interaction assessment suggested heterogeneous user archetypes with distinct change-management needs.

**Conclusions:** Clinical AI deployment should be managed as an organizational learning process rather than as a purely technical implementation. The relevant management object is the deployed sociotechnical system, including workflow, governance architecture, feedback loops, ethics, and user interaction. We propose a Governance-Aware AI Management toolbox that helps administrators and digital health officers distinguish solvable implementation-design uncertainty from true lack of clinical value, and supports staged, evidence-based decisions about scaling, investment, and definitive evaluation.

## 1. Introduction

Health care organizations are navigating an era of AI-enabled uncertainty that reaches beyond individual clinical decisions and into the strategic and operational core of hospital management. As artificial intelligence permeates clinical documentation, resource allocation, reimbursement, vendor ecosystems, workforce planning, and governance structures, executives and digital health officers must decide which systems to build, buy, monitor, scale, or discontinue under incomplete evidence (1–3). The resulting uncertainty is not only statistical or technical. It is organizational: it concerns responsibility, workflow consequences, regulatory exposure, interoperability, internal capability, and whether a pilot generates evidence that is useful for strategic decision-making.

This challenge is particularly visible in hospitals deploying AI-based clinical decision support systems (CDSSs). Such systems are frequently introduced before their optimal form, workflow integration, governance model, and operational role are fully stabilized (4–6). Traditional retrospective model-performance metrics remain necessary, but they do not answer whether a tool can be governed, maintained, trusted, integrated into routine work, or scaled. Conversely, premature outcome-oriented trials may yield negative results because the deployed system is immature rather than because the clinical concept lacks value (7,8).

Critical care provides a relevant empirical setting for studying these management challenges. Intensive care units (ICU) operate under continuous capacity pressure, and length-of-stay (LoS) prediction has direct implications for bed planning, elective surgery scheduling, downstream ward capacity, and staffing. Machine-learning models for ICU LoS and other critical-care prediction tasks have shown promising retrospective performance (9–18), but translation into sustained bedside utility remains limited (6,13,14,19,20). Models can lose performance in live data streams (21–25), but deployment failures are often driven by workflow misfit, lack of calibrated user reliance, insufficient governance infrastructure, data-protection constraints, and unclear accountability (6,26–35).

For hospital leaders, the central question is therefore not only whether an algorithm predicts accurately. It is how uncertainty created by the algorithm, the infrastructure, the users, and the regulatory environment can be managed. In European hospitals, this question is shaped by the General Data Protection Regulation (GDPR), national data-protection law, institutional IT-security policies, the EU Medical Device Regulation (MDR), and the EU AI Act (36–39). These frameworks may force architectural decisions such as pseudonymization, separation from production hospital information systems (HIS), or offline deployment. Such decisions are often framed as compliance requirements, but they also reshape workflow burden, perceived reliability, user trust, and sustainability.

This study uses the KISIK project (“KI-basierte Prognose- und Optimierungsverfahren in Assistenzsystemen zur effektiven und effizienten Steuerung der Intensivkapazität in deutschen Krankenhäusern”) as an empirical case. In KISIK, a machine-learning-based CDSS predicting ICU LoS was prospectively deployed in a surgical ICU and embedded into routine clinical work. The objective was not to claim definitive clinical effectiveness. Instead, the deployment was used to examine which forms of management-relevant uncertainty become visible when a clinical AI system is evaluated as a sociotechnical intervention under governance-constrained real-world conditions.

The aims of this study were: first, to describe how workflow, governance, live benchmarking, embedded ethics, and human-AI interaction shaped a prospective ICU AI deployment; and second, to derive a transferable Governance-Aware AI Management (GAIM) Toolbox for administrators and digital health officers deciding whether and how to scale early-stage clinical AI systems. The contribution is empirical and managerial rather than algorithmic: we provide original prospective deployment data to show how clinical AI pilots can be structured as organizational learning experiments.

## 2. Related Work

### AI-enabled uncertainty as a strategic governance challenge

Health care organizations face mounting pressure to govern AI systems that promise operational gains while introducing new sources of institutional uncertainty (1–3). A scoping review of leadership for AI transformation in health care found that successful adoption requires multidimensional leadership that balances technological opportunities with stakeholder needs and adapts to evolving regulatory and organizational contexts (3). Systematic reviews of AI governance models further suggest that existing frameworks often remain principle-based, fragmented, and insufficiently grounded in empirical deployment evidence (2).

### Learning health systems and continuous AI improvement

The learning health system concept offers a useful lens for clinical AI management because it emphasizes continuous cycles of evidence generation, stakeholder engagement, transparency, equity, and accountability (40). In the AI era, these principles require active feedback loops, performance monitoring, iterative refinement, and mechanisms to learn from real-world use rather than from retrospective validation alone (41). Early clinical AI deployments can therefore be designed as structured organizational learning experiments that build institutional capability before wider rollout.

### Governance under European regulatory uncertainty

European clinical AI deployments are shaped by layered regulatory and institutional requirements, including GDPR, national data-protection provisions, MDR, and the EU AI Act (36–39). These requirements do not merely constrain deployment from outside. They influence architecture, data flows, human oversight, monitoring, and documentation. For administrators, this creates a strategic question: how can a hospital build AI governance that is compliant, operationally feasible, and capable of supporting continuous improvement?

### From pilot evaluation to management evidence

Reporting and evaluation guidance such as TRIPOD+AI and DECIDE-AI has improved transparency around model reporting and early-stage clinical evaluation (8,42). Implementation-oriented frameworks increasingly address workflow integration, human factors, governance, sustainability, and long-term impact (43–48). Figure 1 gives an overview of the existing frameworks. However, practical empirical examples showing how these dimensions can be operationalized simultaneously in resource-realistic hospital deployments remain limited. This study addresses that gap by translating a prospective ICU CDSS deployment into management-relevant evidence for strategic decision-making under AI-enabled uncertainty.

**Figure 1.**
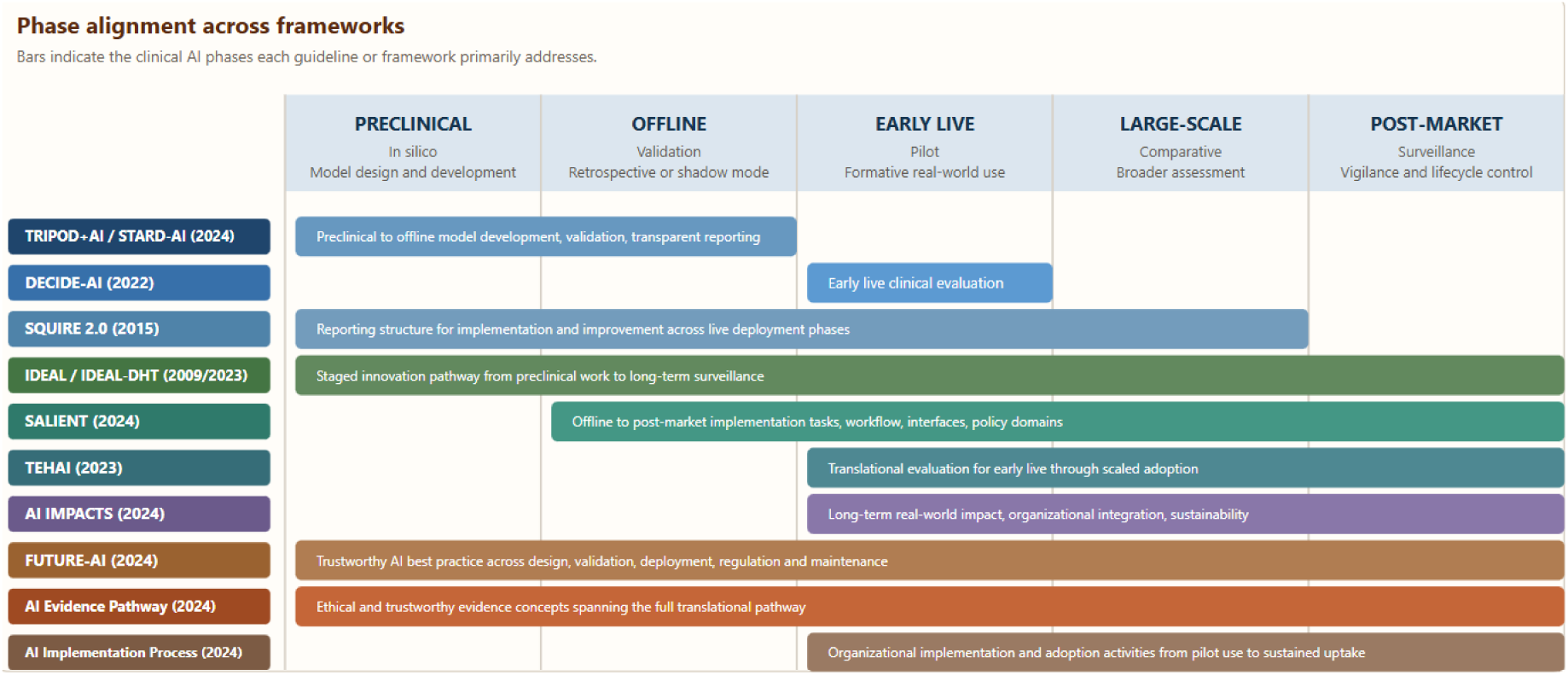
Phase alignment across selected AI and digital health evaluation frameworks.

## 3. Methods

### 3.1 Study Design and Management Orientation

We conducted a prospective, single-center mixed-methods implementation evaluation of an ML-based CDSS for ICU LoS prediction. The study was designed to generate both empirical deployment findings and management-relevant lessons for early-stage clinical AI governance. It was not designed as a definitive clinical effectiveness trial. Instead, it examined whether the deployed sociotechnical system could be integrated into routine ICU work, benchmarked against bedside clinical judgment, iteratively refined, governed under institutional data-protection constraints, and evaluated with respect to ethics and human-AI interaction.

Analyses were descriptive and hypothesis-generating. The management question was whether the pilot produced evidence that could reduce uncertainty for future decisions about scaling, infrastructure investment, governance design, and definitive evaluation.

### 3.2 Setting

The CDSS was deployed in a surgical ICU at a German university hospital from July 1, 2025, to December 1, 2025. The unit operates under continuous bed-pressure conditions and therefore represents a clinically meaningful environment for testing an ICU LoS support tool. Deployment was embedded in routine work rather than in an isolated simulation.

All consecutive ICU stays with an actual ICU LoS longer than 48 hours were eligible. The study was approved by the LMU Munich Institutional Review Board (Project No. 24-0336) and registered with the German Clinical Trials Register (DRKS00037851).

### 3.3 Evaluation Object

The empirical case was a research-prototype CDSS designed to support ICU bed planning by estimating expected ICU LoS from routinely collected hospital and electronic health record data. The underlying model was developed retrospectively using a Machine-Learning Algorithm (HistGradientBoostingRegressor) and subsequently operationalized as a ward-facing application. Technical details of model development and validation are reported in Multimedia Appendix 1.

For this study, the evaluation object was not the prediction algorithm alone. The relevant object was the deployed sociotechnical system, including the model, user interface, data pipeline, governance architecture, explainability layer, onboarding, user interaction, and benchmarking workflow. This distinction was central to the management interpretation: the question was whether the organization could safely and usefully operate the system under real constraints, not whether an isolated model performed well in retrospective data.

#### 3.3.1 Deployment Architecture

The deployed CDSS followed a governance-compliant offline architecture. Patient data were extracted and pseudonymized through the local medical data integration pathway, processed on premises, and transferred to air-gapped ward laptops. The patient roster and model outputs were refreshed once daily. This architecture enabled prospective use in routine care while avoiding direct write access to the production HIS. Two interface iterations were implemented. Version 1 displayed a single LoS estimate; version 2 added an explainability layer based on SHAP values.

#### 3.3.2 Users and Stakeholders

The primary CDSS user group was resident physicians. Experienced consultant physicians used a separate input form to record blinded independent bedside estimates as clinical benchmarks.

Secondary stakeholders included local informatics staff, software developers, governance and data-integration personnel, and the embedded ethics team. Their involvement was necessary because the deployed intervention depended not only on model performance but also on data access, security approval, workflow design, accountability framing, and communication with users.

### 3.4 Evaluation Domains and Outcomes

The evaluation integrated five domains that were prospectively relevant for managing AI-enabled uncertainty:

- **Workflow integration and operational adoption:** clinician roundtables were conducted before deployment to identify realistic decision points, feasible user interactions, and documentation burden.
- **Governance architecture and data-protection effects:** governance and data-protection requirements were mapped as part of the evaluation, including pseudonymization, IT security, separation from production HIS infrastructure, and restrictions on real-time connectivity.
- **Live bedside benchmarking and continuous learning:** resident physicians used the AI-supported workflow, whereas consultant physicians remained blinded to CDSS output and provided independent bedside estimates. This created a contemporaneous real-world comparator and enabled iterative refinement.
- **Embedded ethics and accountability design:** an ethics expert was involved from the outset using an Embedded Ethics and Social Sciences approach to translate ethical concerns into operational design decisions (49–51). Ethical considerations were continuously embedded across both the development and implementation phases. The ethicist actively participated in stakeholder engagement activities and project meetings, critically examining the ethical challenges associated with deploying AI in the ICU.
- **Human-AI interaction and change-management needs:** human-AI interaction was assessed using the Psychological Assessment of AI-based Decision Support Systems (PAAI) instrument among 11 physicians. Exploratory k-means clustering of normalized response profiles was used descriptively to identify provisional user archetypes (52).

The main empirical outcomes included usage proportions among consultants and resident physicians, completeness of consultant benchmark capture, CDSS mean absolute error before and after the first iterative update, frequency of outdated case-list entries, ethics-derived design outputs, and exploratory PAAI-based user archetypes.

### 3.5 Statistical and Descriptive Analysis

Descriptive statistics were used to summarize usage, benchmark capture, prediction error, and governance-related workflow effects. Prediction error was summarized using mean absolute error. PAAI responses were normalized and explored using k-means clustering to characterize provisional user profiles. No causal inference regarding patient outcomes or system-level effectiveness was intended.

## 4. Results

### 4.1 Workflow Integration and Operational Adoption

Pre-deployment clinician roundtables identified ICU-specific workflow constraints and natural decision points. The resulting low-burden “glance-and-judge” interface enabled access to predictions and documentation within existing routines. The co-design process prevented development of a more complex dashboard that stakeholders considered unlikely to fit the ICU work environment.

The evaluation included 267 consecutive ICU stays. Figure 2 shows the workflow integration of the CDSS. The CDSS was technically deployable under routine ICU conditions despite the absence of direct HIS integration. Routine use was achieved among both clinician groups, indicating that the basic workflow concept was feasible. From a management perspective, this finding showed that adoption capacity depended on aligning the tool with existing temporal and role-specific routines rather than on technical availability alone.

**Figure 2.**
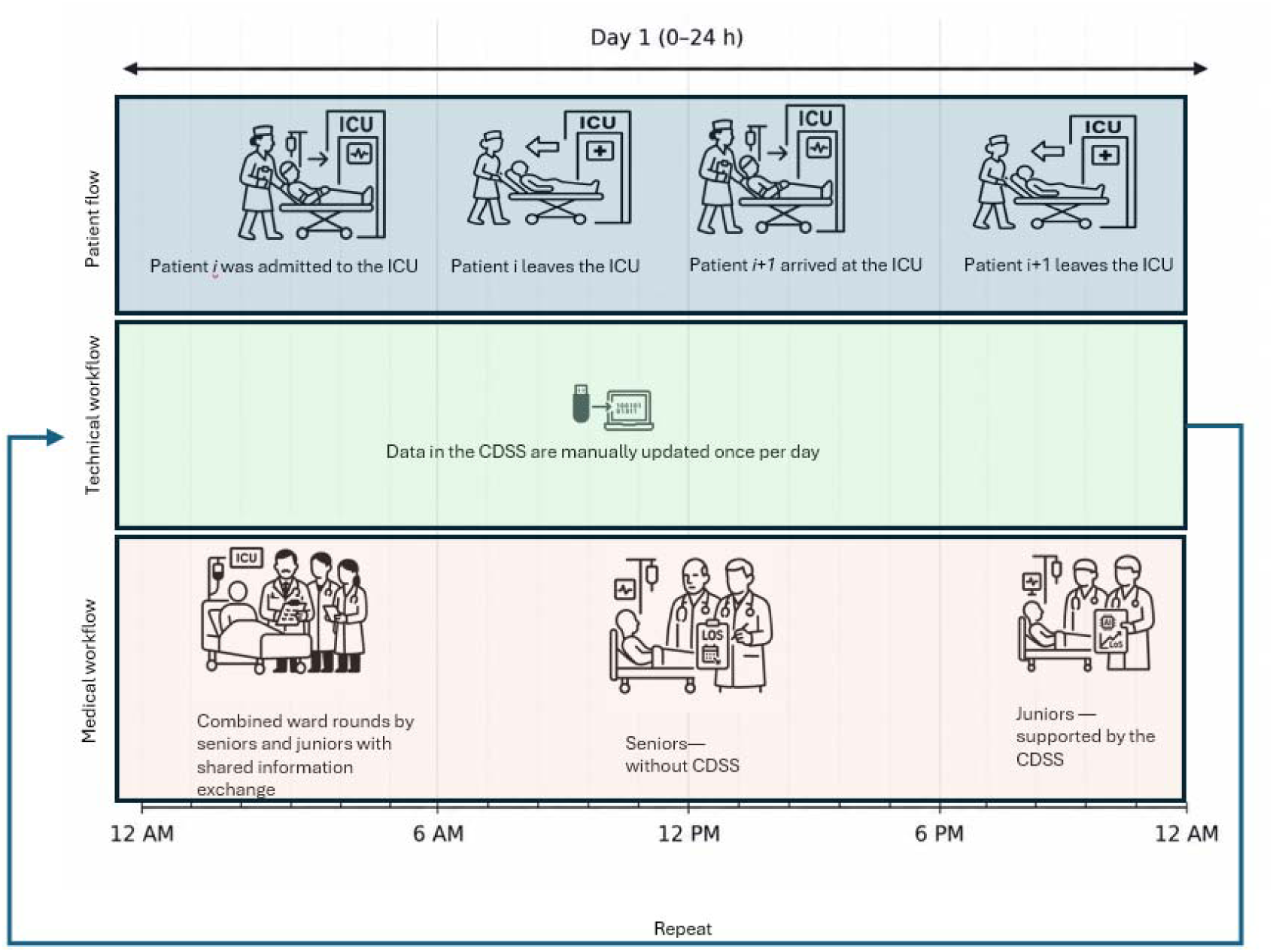
Workflow integration of the clinical decision support system (CDSS) into routine ICU work.

A usage proportion of 72% (191/267) was achieved among consultants and 61% (162/267) among resident physicians. Usage patterns showed that the system reached its intended environment but did so asymmetrically across professional roles and time windows. Residents predominantly interacted with the CDSS during night-time work, whereas consultants used the benchmarking workflow during daytime planning contexts. This workflow asymmetry is relevant for interpreting adoption, benchmarking, and change-management findings. Figure 3 visualizes this context.

**Figure 3a.**
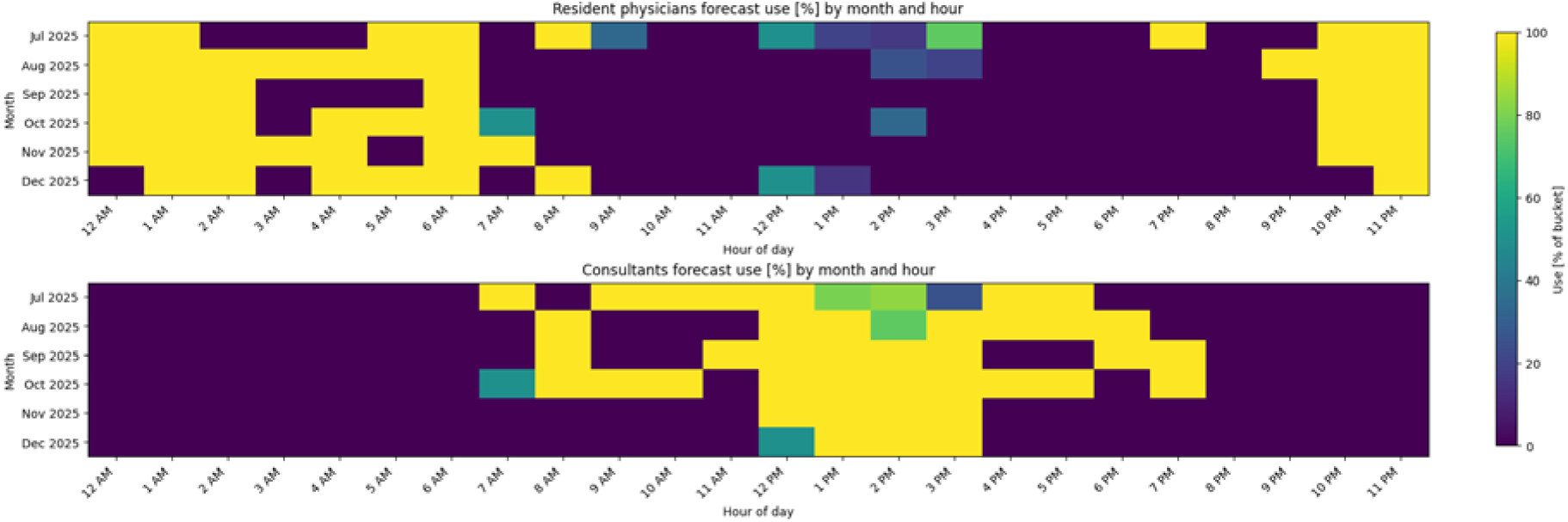
Temporal usage heatmap of resident and consultant interaction with the CDSS and benchmarking workflow.

**Figure 3b.**
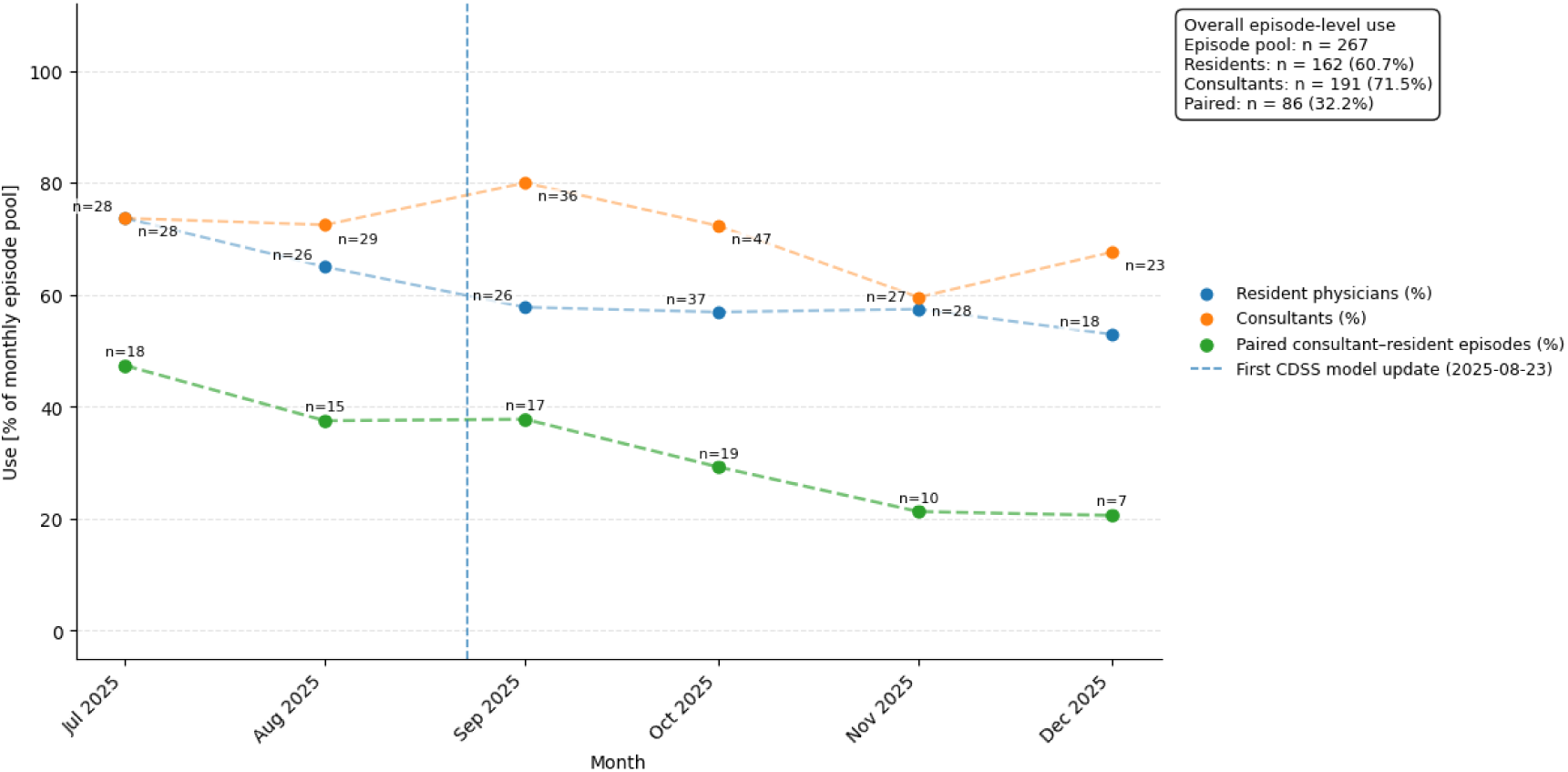
Monthly benchmark capture and usage trends during the implementation period.

### 4.2 Governance Architecture and Infrastructure Consequences

Governance and IT-security requirements shaped the deployment architecture. Because real-time HIS connectivity was not possible, the CDSS used an air-gapped architecture with once-daily updates.

Figure 4 illustrates the deployment architecture. This enabled governance-compliant deployment but introduced potential mismatch between CDSS case lists and the live ward census.

**Figure 4.**
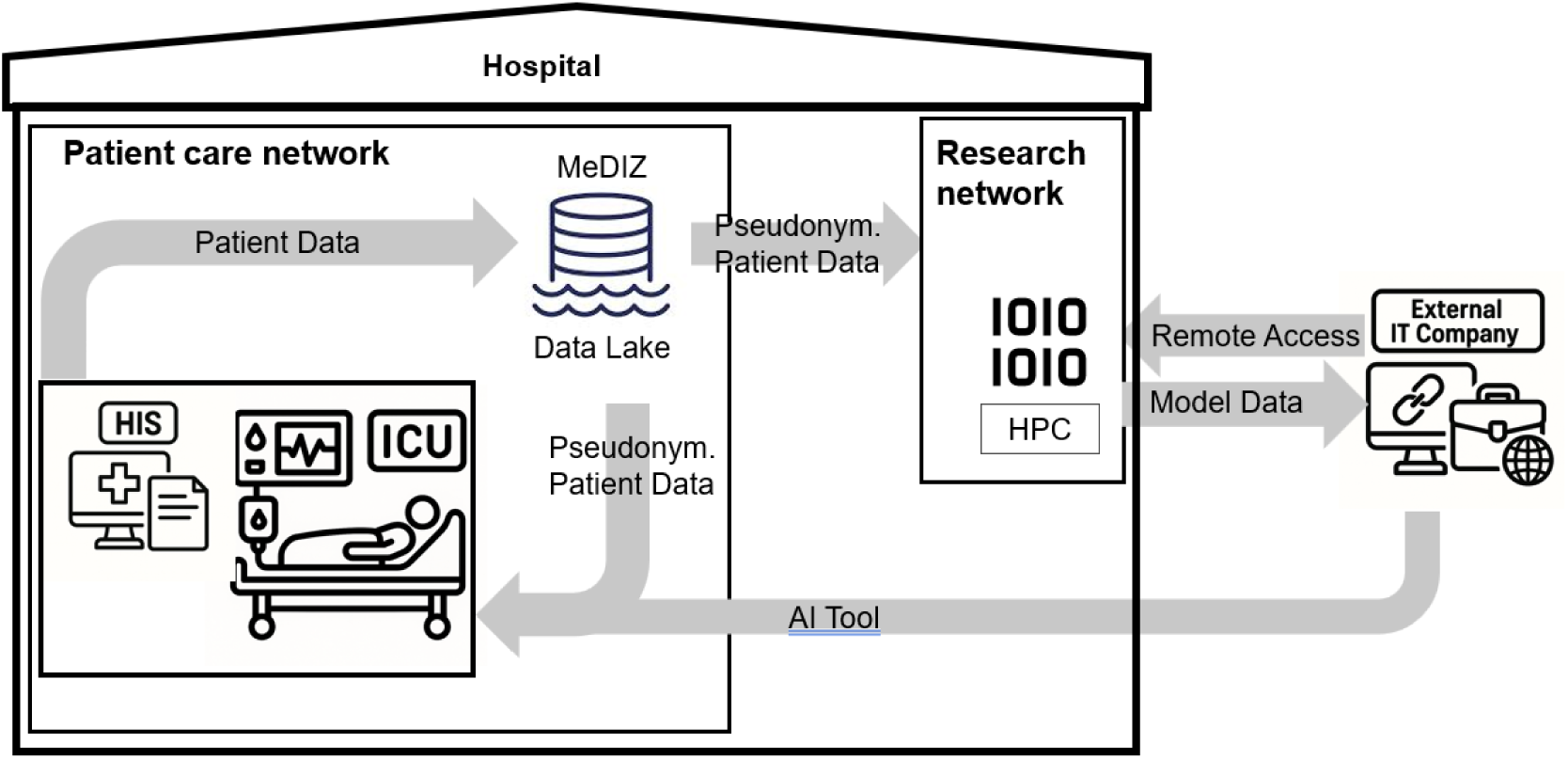
Deployment architecture under governance constraints. HIS: hospital information system; MeDIZ: medical data integration center.

During the analyzed implementation period, outdated case-list entries occurred in 4 of 148 benchmarked stays (2.7%). Numerically, the rate was modest. Operationally, however, the issue was salient because even a small number of outdated records can reduce trust in a time-critical environment. Clinicians compensated by manually cross-checking the live ward census before acting on the CDSS roster. This transferred coordination work from infrastructure to frontline users.

For administrators, this result illustrates that governance architecture is not only a compliance decision. It creates operational latency, hidden work, and potential trust effects that should be considered when comparing offline, semi-integrated, and fully integrated deployment pathways.

### 4.3 Embedded Ethics and Accountability Design

The Embedded Ethics and Social Sciences approach translated ethical and social questions into concrete development and implementation decisions (51). A systematic literature review identified ethical issues reported in comparable AI and critical-care projects. Semi-structured guided interviews were conducted with all KISIK consortium members, including software developers, clinicians, and project managers. Across 12 interviews lasting between 45 and 90 minutes, ethical and social challenges were explored in depth. Interviews were audio-recorded, transcribed verbatim, and analyzed in MAXQDA using thematic analysis.

Findings were anonymized and shared with the broader project team. This process identified ethical strengths and vulnerabilities and informed subsequent development decisions. Explainability was examined not solely as a technical feature but as an accountability mechanism: what function should explanations serve for clinicians in practice, and how should they support critical disagreement with algorithmic outputs? The CDSS was explicitly framed as decision support rather than decision automation. Interface wording, onboarding, and accountability framing were adjusted to preserve clinicians’ permission to disagree with AI-generated outputs.

Ethics therefore functioned not as a one-time approval procedure but as a formative governance mechanism embedded throughout the project lifecycle. For management, this produced actionable design outputs on responsibility, automation bias, explainability, and communication standards.

### 4.4 Live Benchmarking as an Organizational Learning Loop

Prospective consultant benchmarking provided a contemporaneous real-world comparator for CDSS predictions. This enabled the evaluation to compare AI predictions with bedside clinical judgment under routine conditions rather than with retrospective benchmarks alone. After the first iterative model update, CDSS mean absolute error decreased from 5.95 to 4.12 days. This finding is interpreted as an exploratory implementation result, not definitive evidence of clinical benefit. Figure 5 shows that approach at an exploratory level.

**Figure 5.**
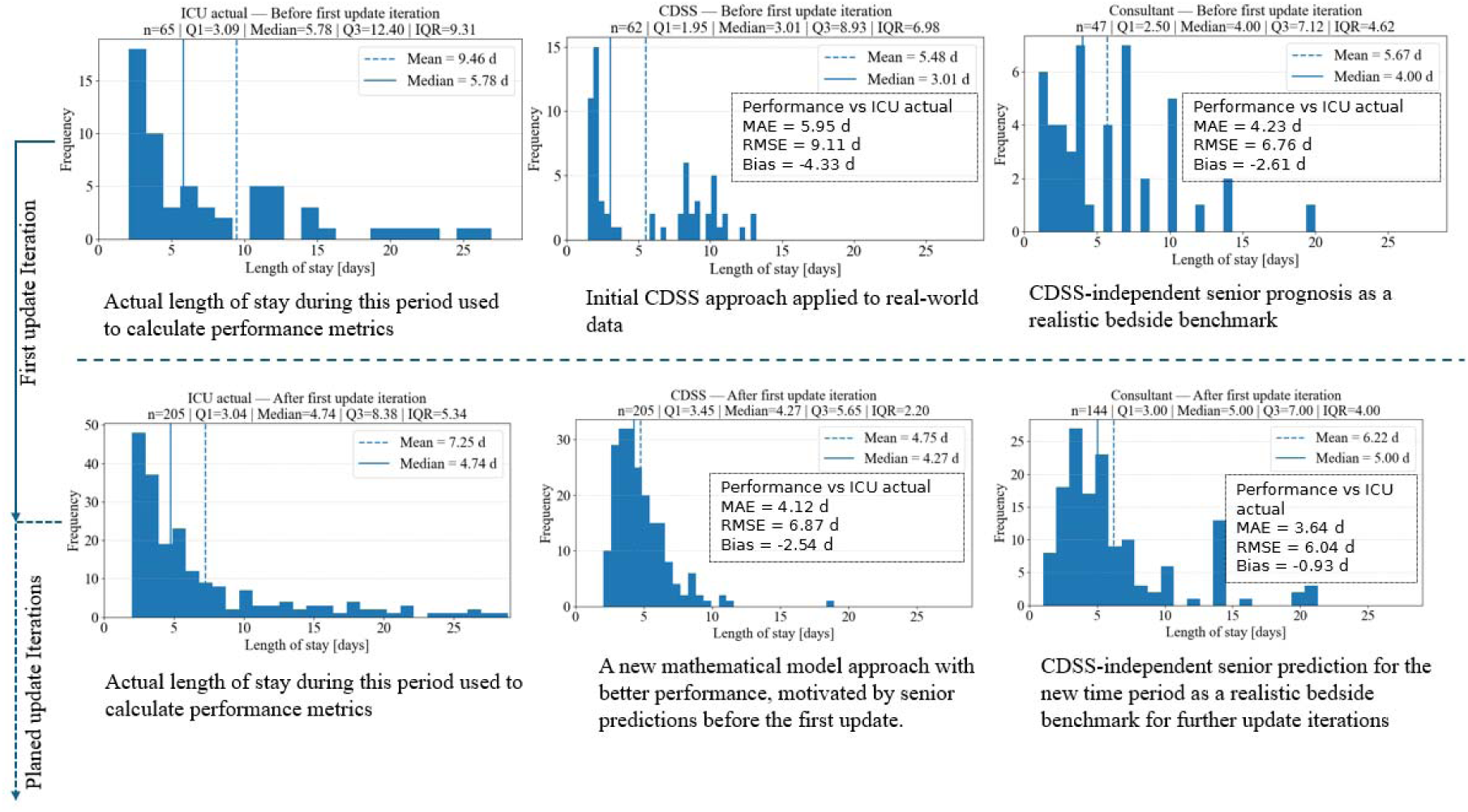
Live bedside benchmarking and illustrative iterative model update.

From a management perspective, the important finding was not the magnitude of improvement alone but the presence of a feedback loop. Live deployment identified performance and plausibility gaps, guided targeted updates, and enabled reassessment under similar operational conditions. This converted an AI pilot from a static test into a learning mechanism for the clinical unit and the organization.

### 4.5 User Archetypes and Change-Management Implications

PAAI assessment among 11 physicians suggested heterogeneous interaction patterns. Exploratory clustering identified three provisional user archetypes (Figure 6). Despite the small sample, these patterns illustrate why average acceptance scores are insufficient for managing clinical AI adoption.

**Figure 6.**
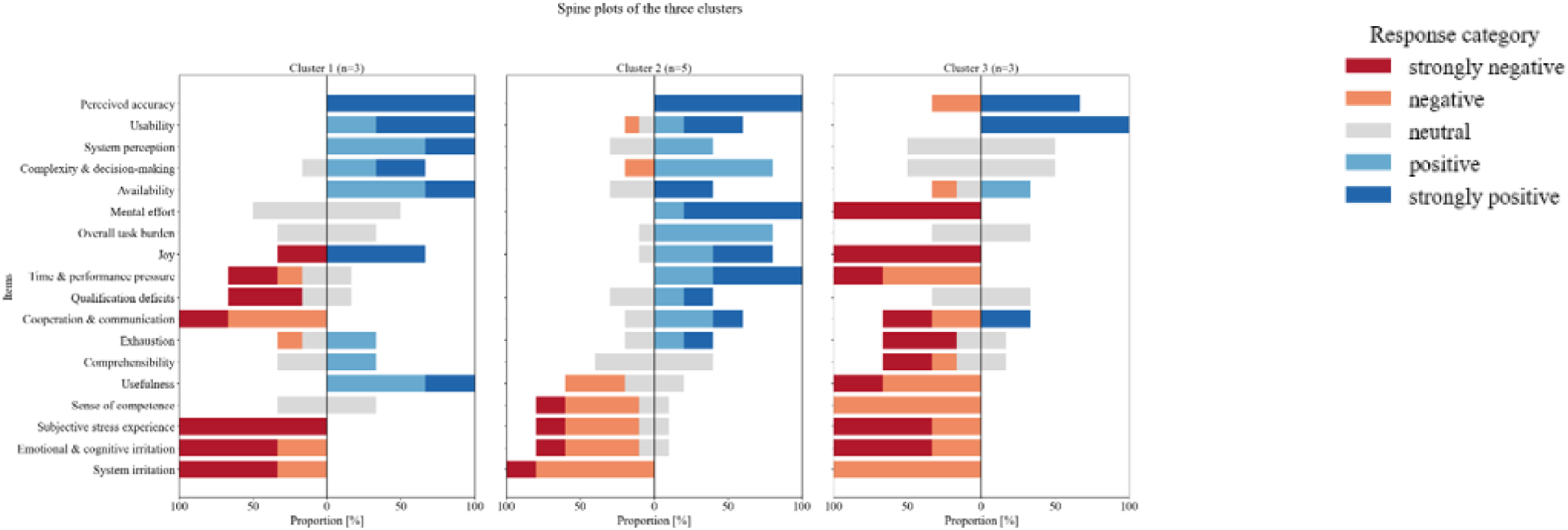
Exploratory PAAI-based user archetypes among clinicians.

Cluster 1 (pragmatic users) perceived clear value in AI-supported estimation and welcomed additional explanatory depth. Cluster 2 (enthusiastic adopters) showed workable acceptance, with perceived benefit depending on efficiency and clinical fit. Cluster 3 (time-pressured skeptics) were not categorically opposed to AI, but perceived interaction costs as outweighing practical benefit. These patterns were not predictable based on seniority alone.

The finding suggests that administrators should not assume uniform adoption after onboarding. Clinical AI implementation requires targeted change-management strategies that distinguish nonuse, appropriate disagreement, lack of understanding, and calibrated reliance.

### 4.6 Governance-Aware AI Management Toolbox

Based on the empirical findings, we synthesized the Governance-Aware AI Management Toolbox (GAIM-Toolbox) for early-stage clinical AI deployment. GAIM-Toolbox treats the deployed sociotechnical system as the management object and organizes uncertainty into five connected domains: workflow and operational adoption, governance architecture and infrastructure consequences, continuous learning through live benchmarking, embedded ethics and accountability design, and human-AI interaction with change-management needs. Figure 7 illustrates these domains as a staged management toolbox, and Table 1 summarizes the empirical uncertainty surfaced in KISIK and the corresponding strategic implications.

**Figure 7.**
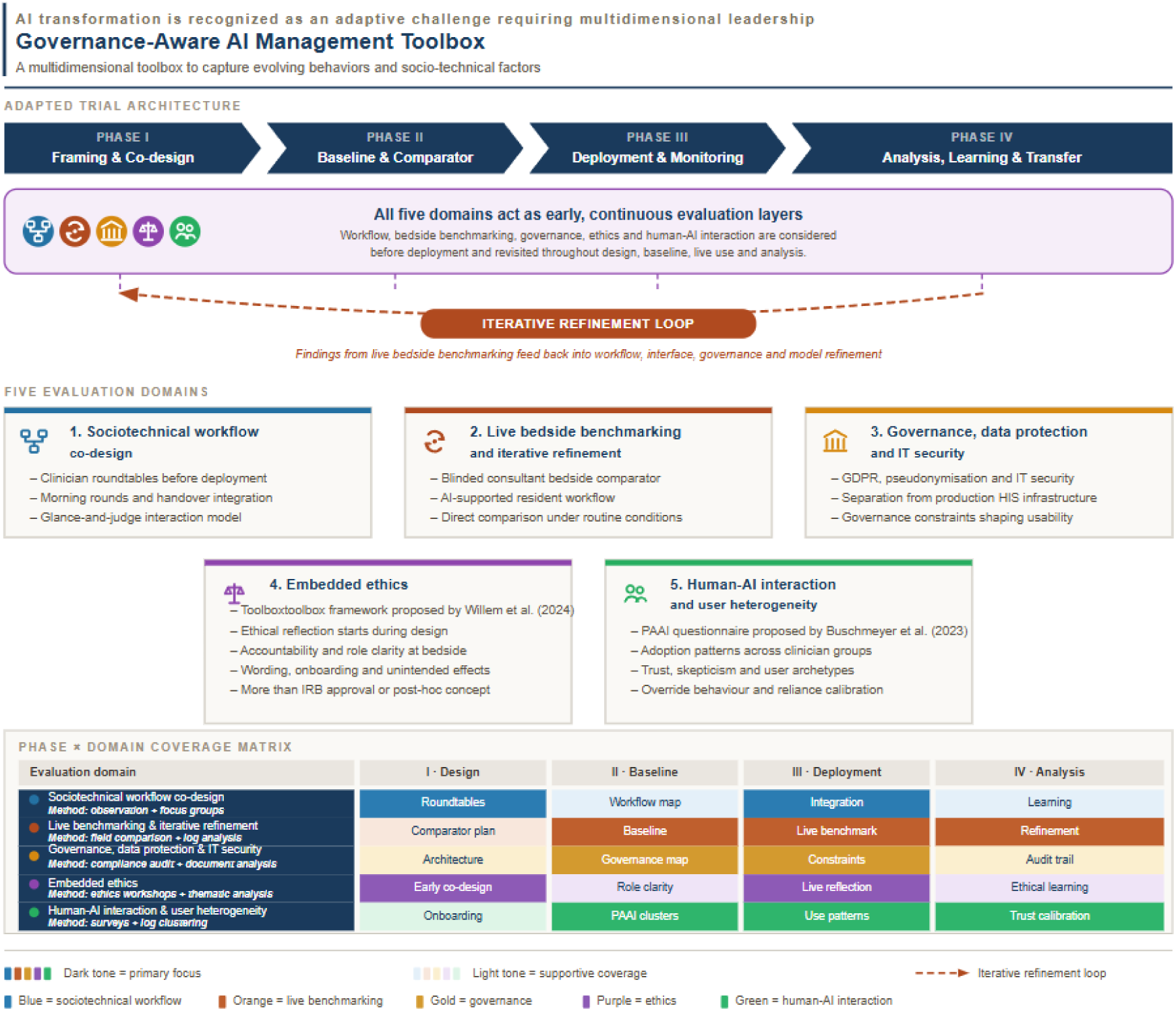
Governance-Aware AI Management Toolbox (GAIM-Toolbox) derived from the KISIK deployment.

**Table 1.** Governance-Aware AI Management Toolbox: uncertainty domains, empirical signals, and strategic implications.

| Management domain | Empirical uncertainty surfaced in KISIK | Strategic implication for administrators and digital health officers |
| --- | --- | --- |
| Workflow and operational adoption | Technical availability did not determine use; adoption depended on shift timing, role-specific routines, and low-burden interaction design. | Co-design should precede deployment. Usage data should be interpreted in relation to workflow context, not as a simple binary marker of acceptance. |
| Governance architecture and infrastructure consequences | Data-protection and IT-security requirements forced an air-gapped architecture, creating latency and hidden frontline cross-checking work. | Governance choices should be treated as strategic design variables with measurable operational costs, not only as compliance prerequisites. |
| Continuous learning through live benchmarking | Retrospective validation alone could not show how the CDSS compared with live clinical judgment or where refinement was needed. | AI pilots should include feedback loops that benchmark performance in context and support staged refinement before scale-up. |
| Embedded ethics and accountability design | Abstract ethical principles required translation into concrete wording, onboarding, explainability, and responsibility arrangements. | Ethics should be embedded as a formative governance function that shapes accountability, calibrated reliance, and permission to disagree. |
| Human-AI interaction and change management | PAAI profiles suggested heterogeneous adoption needs that were not explained by seniority alone. | Implementation plans should include targeted change-management strategies for different user archetypes and distinguish adoption from meaningful use. |

## 5. Discussion

### 5.1 Principal Findings

This prospective mixed-methods ICU deployment shows that clinical AI pilots can generate management-relevant evidence when the deployed sociotechnical system is evaluated rather than the algorithm alone. The principal finding is that AI-enabled uncertainty became visible across five connected domains: workflow, governance architecture, continuous learning, embedded ethics, and human-AI interaction. Several findings would have remained invisible in a conventional performance-focused evaluation, including governance-driven latency, hidden coordination work, role- and time-dependent use, user archetype heterogeneity, and the value of live benchmarking as an organizational learning loop.

The study does not claim that the CDSS improved patient outcomes or ICU capacity management. Its contribution is more specific: it demonstrates how an early-stage AI deployment can help administrators distinguish between uncertainty that is intrinsic to the clinical prediction task and uncertainty that results from implementation design, governance architecture, workflow misfit, or change-management needs.

### 5.2 Strategic Implications for Health Care Management

First, governance architecture should be treated as a strategic design variable. In KISIK, the air-gapped architecture enabled compliance and reduced risk, but it also created operational latency and shifted cross-checking work to frontline clinicians. Similar trade-offs are likely whenever hospitals decide between research-prototype deployment, semi-integrated pilots, vendor-mediated integration, and full production-system connectivity. Administrators should therefore quantify not only direct technology costs but also hidden coordination work, trust effects, update frequency, and responsibility boundaries.

Second, clinical AI pilots should be designed as structured learning experiments (53). The live benchmarking component allowed the organization to observe AI performance against bedside clinical judgment under routine conditions, identify gaps, and refine the model. This operationalizes learning health system principles at unit level: data from real-world use are converted into feedback for improvement, governance learning, and future investment decisions (40,41).

Third, change management must account for heterogeneous user interaction. The PAAI-based profiles suggested that clinicians differed in perceived usefulness, comprehensibility, irritation, and workload implications. These differences were not reducible to seniority. Administrators should therefore avoid implementation strategies that assume a homogeneous workforce. Targeted onboarding, explainability depth, feedback channels, and role-specific use cases may be needed to achieve calibrated reliance rather than superficial adoption.

Fourth, embedded ethics can function as a management tool. By translating concerns about automation bias, responsibility, explainability, and permission to disagree into design and onboarding decisions, the ethics component helped align the CDSS with professional accountability. This is relevant for AI governance because hospitals must not only comply with external rules but also create internal norms for how clinicians should question, document, override, and learn from AI recommendations.

### 5.3 Relationship to Learning Health Systems and Adaptive Governance

The KISIK deployment illustrates how early-stage clinical AI can be embedded within a learning health system logic. Rather than freezing the intervention for immediate definitive testing, the pilot created feedback loops across clinical use, benchmarking, ethics, user feedback, and governance mapping. This does not imply unregulated trial-and-error. The learning process itself requires oversight, documentation, accountability, and clear boundaries for model updates and clinical use.

Adaptive governance is therefore central. Administrators need mechanisms for deciding when a prototype remains a research tool, when it may be integrated into routine operations, when it requires regulatory classification or conformity assessment, and when performance or workflow signals should trigger redesign. GAIM provides a practical structure for organizing these decisions across technical, organizational, ethical, and human-factor dimensions.

### 5.4 Comparison With Prior Work

Our findings are consistent with prior work showing that clinical AI translation depends on more than model performance(13,14,20). They also align with implementation literature emphasizing workflow fit, stakeholder engagement, and human factors (28–31). The contribution of this study is to connect these domains explicitly to strategic management under AI-enabled uncertainty. Existing guidance such as TRIPOD+AI and DECIDE-AI supports transparent reporting and early-stage live evaluation (8,42), while broader frameworks address governance, impact, sustainability, and trustworthy deployment (43–48). GAIM adds an empirical management layer by showing how these dimensions interact in one governance-constrained hospital deployment (Figure 8).

**Figure 8.**
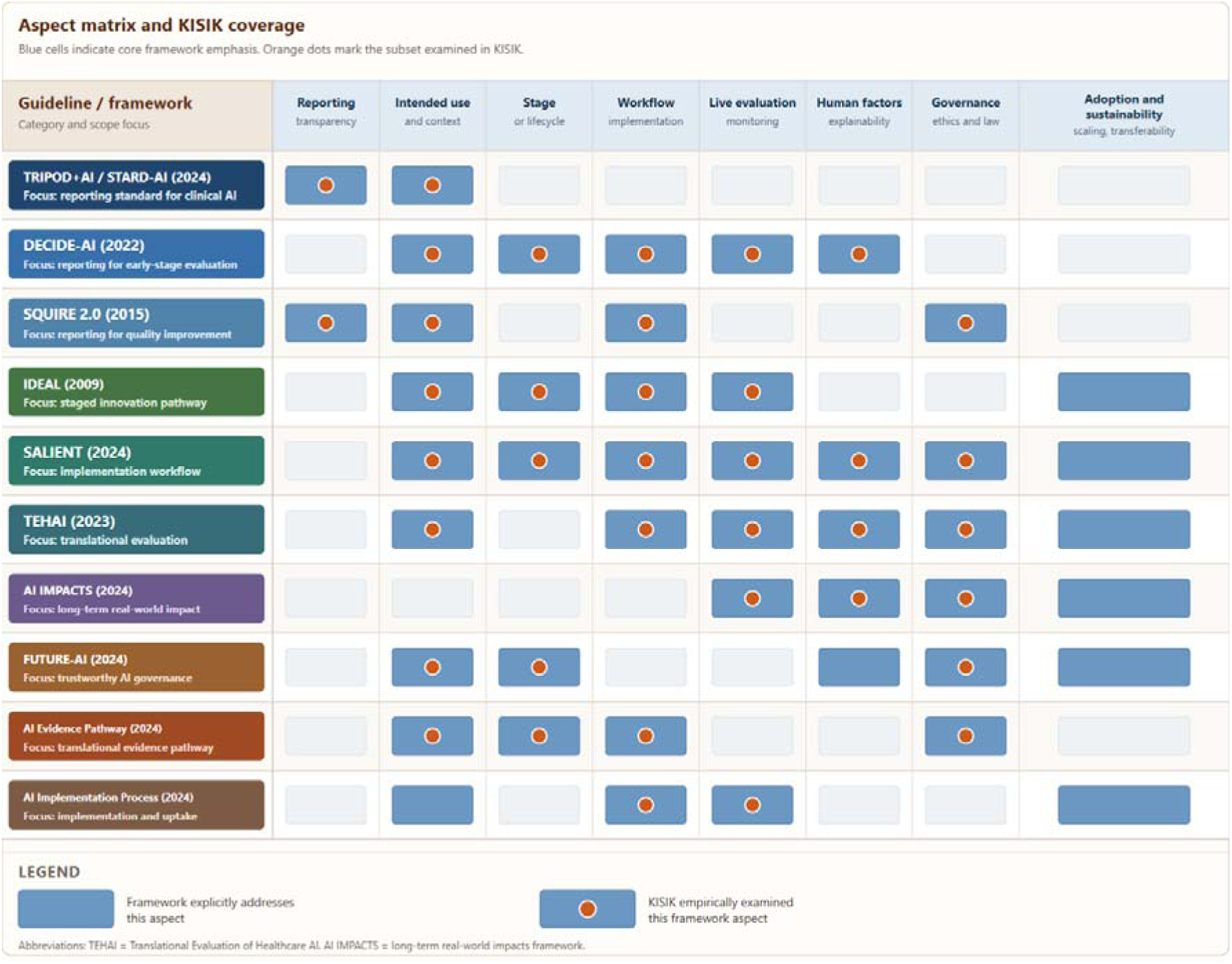
Coverage of selected framework aspects by the GAIM-Toolbox.

The study also extends discussions of European AI governance by showing how regulatory and institutional constraints become operational phenomena. Data protection and IT-security rules did not merely determine what was legally permissible; they shaped the actual system users experienced, including data latency and extra work. This finding is important for hospitals comparing internal development, vendor procurement, cloud infrastructure, or hybrid architecture.

### 5.5 Implications for Definitive Evaluation and Scaling

Definitive trials and outcome evaluations remain necessary to determine clinical and operational effectiveness. However, this case suggests that trials should be timed after key implementation uncertainties have been characterized. A negative trial conducted before resolving workflow, governance, performance-monitoring, or user-interaction issues may be difficult to interpret because poor outcomes could reflect implementation-design failure rather than lack of clinical value.

Conversely, formative evaluation without a path toward outcome assessment risks becoming an endless pilot. GAIM is intended to support staged decisions: redesign, discontinue, maintain as a research prototype, scale within a controlled governance pathway, or proceed to outcome-oriented testing.

### 5.6 Limitations

This study has several limitations. It was conducted at a single German university hospital in a surgical ICU and focused on an ICU LoS prediction CDSS. The specific model, data pipeline, patient population, and governance architecture may not generalize directly to other settings. Consultant benchmark capture was incomplete, and the PAAI sample was small (n=11) and analyzed descriptively. The study did not evaluate downstream patient outcomes or operational outcomes such as discharge timing, cancellation rates, ward throughput, or economic value. The outdated case-list issue was quantified descriptively but not assessed through formal workload measurement.

These limitations define the scope of the contribution. The study does not claim definitive clinical effectiveness of the CDSS. It provides a management-oriented case study and a Toolbox for generating implementation and governance evidence before scale-up or definitive effectiveness testing.

### 5.7 Future Work

Future work should test GAIM across multiple digital health interventions, clinical settings, and governance architectures. For clinical AI systems, next-step evaluation should include improved data synchronization, formal drift and calibration monitoring, expanded workflow and workload measurement, patient- and system-level outcome assessment, economic analysis, and multi-site testing. Comparative studies should examine whether structured governance-aware pilot evaluation improves scaling decisions, reduces pilot failure, and strengthens later definitive trials.

## 6. Conclusions

This prospective ICU deployment illustrates that clinical AI management must address more than model performance. In real-world deployment, uncertainty arose from workflow integration, governance architecture, data latency, clinician interaction, accountability framing, and the capacity to learn from live use. The relevant management object was the deployed sociotechnical system, not the algorithm alone.

We propose the Governance-Aware AI Management Toolbox as a practical structure for administrators and digital health officers navigating early-stage clinical AI deployment. The toolbox supports staged decisions about redesign, monitoring, scaling, and definitive evaluation. In European hospital settings in particular, governance should be treated as a primary design and evaluation variable from the outset. Early AI pilots should be used to build organizational learning capacity, quantify hidden implementation costs, and distinguish solvable deployment uncertainty from true lack of clinical value.

## Data Availability Statement

For data access inquiries, please contact the corresponding author. Access to patient-level data is restricted because the data derive from routinely collected clinical information and are subject to institutional data-protection requirements.

## Ethics Statement

The study was performed in accordance with the Declaration of Helsinki and was reviewed and approved by the LMU Munich Institutional Review Board (Project No. 24-0336; registry: DRKS00037851). Individual patient consent was not obtained because the analysis was based on routinely collected clinical data generated during standard care and processed in pseudonymized form. The study did not involve study-specific diagnostic or therapeutic interventions in patients.

## Author Contributions

Alexander Althammer: Conceptualization; Methodology; Investigation; Data curation; Formal analysis; Project administration; Writing – original draft; Writing – review & editing.

Anton Hummel: Software; Data curation; Methodology; Validation; Writing – review & editing.

Jan-Philipp Steghöfer: Software. Felix Reichel: Software.

Jonas Kolonko: Software.

Sarah Hatfield: Methodology; Investigation; Formal analysis.

Maximilian Fischer: Methodology; Investigation.

Kerstin Schlögl-Flierl: Conceptualization; Methodology; Investigation; Supervision.

Paula Ziethmann: Methodology; Investigation; Formal analysis; Writing – review & editing.

Manfred Weiß: Investigation; Resources; Supervision; Writing – review & editing.

Philipp Simon: Investigation; Resources; Supervision; Writing – review & editing.

Markus Mögerlein: Investigation; Resources; Writing – review & editing.

Eugen Mamtschur: Resources; Data curation; Software.

Oliver Spring: Investigation; Resources; Writing – review & editing.

Sergey Shmygalev: Investigation; Resources; Writing – review & editing.

Natalia Ortmann: Resources; Data curation; Software; Writing – review & editing.

Johannes Raffler: Resources; Data curation; Software; Writing – review & editing.

Christian Hinske: Resources; Data curation.

Jens O. Brunner: Conceptualization; Methodology; Supervision; Funding acquisition; Writing – review & editing.

Axel R. Heller: Conceptualization; Resources; Supervision; Funding acquisition; Writing – review & editing.

Christina C. Bartenschlager: Conceptualization; Methodology; Supervision; Project administration; Funding acquisition; Writing – review & editing.

All authors reviewed and approved the final manuscript.

## Declaration of Generative AI and AI-Assisted Technologies

ChatGPT (OpenAI, GPT-4o) was used for translation and linguistic editing. It was not used to generate scientific conclusions or perform data analysis. All AI-assisted outputs were reviewed and revised by the authors, who take full responsibility for the final content.

## Acknowledgments

We thank the physicians of the surgical and anesthesiology departments at the Faculty of Medicine, University of Augsburg, for participating in this study. We also thank the local informatics, governance, and data-integration teams who supported the deployment.

## Supplementary Material

Supplementary File 1. Technical model-development details

Detailed information on retrospective model development, preprocessing, feature handling, training strategy, model selection, and validation procedures.

## References

1. Tian J, Zhao Z, Tang L, Song Y, Li Y, Jiang N. From Pilot Trap to Institutional Capacity: A Governance Framework for Sustainable Clinical AI Implementation in Health Systems. J Med Internet Res. 2026 May 7;28:e92680–e92680. doi:10.2196/92680

2. Hussein R, Zink A, Ramadan B, Howard FM, Hightower M, Shah S, et al. Advancing healthcare AI governance through a comprehensive maturity model based on systematic review. npj Digit Med. 2026 Feb 11;9(1):236. doi:10.1038/s41746-026-02418-7

3. Sriharan A, Sekercioglu N, Mitchell C, Senkaiahliyan S, Hertelendy A, Porter T, et al. Leadership for AI Transformation in Health Care Organization: Scoping Review. J Med Internet Res. 2024 Aug 14;26:e54556. doi:10.2196/54556

4. Sujan M. Integrating digital health technologies into complex clinical systems. BMJ Health Care Inform. 2023 Oct 13;30(1). doi:10.1136/bmjhci-2023-100885 PubMed PMID: 10.1136/bmjhci-2023-100885.

5. Wittich L, Rödiger H, Rombey T, Brecher AL, Kraft L, Sgraja S, et al. Navigating the complexities of digital health technology implementation: a scoping review of barriers and facilitators. Implement Sci Commun. 2026 Mar 4;7(1):69. doi:10.1186/s43058-026-00892-4

6. van de Sande D, van Genderen ME, Huiskens J, Gommers D, van Bommel J. Moving from bytes to bedside: a systematic review on the use of artificial intelligence in the intensive care unit. Intensive Care Med. 2021;47(7):750–60. doi:10.1007/s00134-021-06446-7 PubMed PMID: 34089064; PubMed Central PMCID: PMC8178026.

7. Rosenthal JT, Beecy A, Sabuncu MR. Rethinking clinical trials for medical AI with dynamic deployments of adaptive systems. npj Digit Med. 2025 May 6;8(1):252. doi:10.1038/s41746-025-01674-3

8. Vasey B, Nagendran M, Campbell B, Clifton DA, Collins GS, Denaxas S, et al. Reporting guideline for the early-stage clinical evaluation of decision support systems driven by artificial intelligence: DECIDE-AI. Nat Med. 2022 May;28(5):924–33. doi:10.1038/s41591-022-01772-9 PubMed PMID: 35585198.

9. Workum JD, Meyfroidt G, Bakker J, Jung C, Tobin JM, Gommers D, et al. AI in critical care: A roadmap to the future. Journal of Critical Care. 2026 Feb;91:155262. doi:10.1016/j.jcrc.2025.155262

10. Cecconi M, Greco M, Balzani E, Aliverti A, Azoulay E, Bignami E, et al. Innovation in Intensive Care: a framework to turn ideas and concepts into actionable solutions [Internet]. 2026 Mar 24. doi:10.17863/CAM.128637

11. Freyer O, Mathias R, Muti HS, Orlovsky H, Buch S, Ostermann M, et al. The regulation of artificial intelligence in intensive care units: from narrow tools to generalist systems. npj Digit Med. 2026 Mar 21;9(1):246. doi:10.1038/s41746-026-02535-3

12. Saqib M, Iftikhar M, Neha F, Karishma F, Mumtaz H. Artificial intelligence in critical illness and its impact on patient care: a comprehensive review. Front Med. 2023 Apr 20;10. doi:10.3389/fmed.2023.1176192

13. Berkhout WEM, van Wijngaarden JJ, Workum JD, van de Sande D, Hilling DE, Jung C, et al. Operationalization of Artificial Intelligence Applications in the Intensive Care Unit: A Systematic Review. JAMA Netw Open. 2025 Jul 23;8(7):e2522866. doi:10.1001/jamanetworkopen.2025.22866

14. Sokol K, Fackler J, Vogt JE. Artificial intelligence should genuinely support clinical reasoning and decision making to bridge the translational gap. npj Digit Med. 2025 Jun 10;8(1):345. doi:10.1038/s41746-025-01725-9

15. Garber S, Okhrin Y. Machine Learning for Intensive Care Unit Length-of-Stay Prediction: A Simulation-Based Approach to Bed Capacity Management. Med Decis Making. 2026 Apr 1;46(3):355–70. doi:10.1177/0272989X251406639

16. Park DJ, Baik SM, Hong KS, Yi H, Lee JG, Lee JM. Development and external validation of an artificial intelligence model for predicting mortality and prolonged ICU stay in postoperative critically ill patients: a retrospective study. World J Emerg Surg. 2025 Oct 15;20(1):79. doi:10.1186/s13017-025-00650-2

17. Goh KH, Wang L, Yeow AYK, Poh H, Li K, Yeow JJL, et al. Artificial intelligence in sepsis early prediction and diagnosis using unstructured data in healthcare. Nat Commun. 2021 Jan 29;12(1):711. doi:10.1038/s41467-021-20910-4

18. Aldhoayan MD, Aljubran Y. Prediction of ICU Patients’ Deterioration Using Machine Learning Techniques. Cureus. 15(5):e38659. doi:10.7759/cureus.38659 PubMed PMID: 37288226; PubMed Central PMCID: PMC10242424.

19. Show us the evidence for the value of medical AI. Nat Med. 2026 Apr;32(4):1163–1163. doi:10.1038/s41591-026-04389-4

20. Van de Sande D, van Genderen M, Braaf H, Gommers D, van Bommel J. Moving towards clinical use of artificial intelligence in intensive care medicine: business as usual? Intensive Care Medicine. 2022 Oct 21;48. doi:10.1007/s00134-022-06910-y

21. Subasri V, Krishnan A, Kore A, Dhalla A, Pandya D, Wang B, et al. Detecting and Remediating Harmful Data Shifts for the Responsible Deployment of Clinical AI Models. JAMA Netw Open. 2025 Jun 4;8(6):e2513685. doi:10.1001/jamanetworkopen.2025.13685 PubMed PMID: 40465297; PubMed Central PMCID: PMC12138723.

22. Schinkel M, Boerman AW, Paranjape K, Wiersinga WJ, Nanayakkara PWB. Detecting changes in the performance of a clinical machine learning tool over time. eBioMedicine. 2023 Oct 2;97:104823. doi:10.1016/j.ebiom.2023.104823 PubMed PMID: 37793210; PubMed Central PMCID: PMC10550508.

23. Davis SE, Greevy RA, Lasko TA, Walsh CG, Matheny ME. Detection of Calibration Drift in Clinical Prediction Models to Inform Model Updating. J Biomed Inform. 2020 Dec;112:103611. doi:10.1016/j.jbi.2020.103611 PubMed PMID: 33157313; PubMed Central PMCID: PMC8627243.

24. Kore A, Abbasi Bavil E, Subasri V, Abdalla M, Fine B, Dolatabadi E, et al. Empirical data drift detection experiments on real-world medical imaging data. Nat Commun. 2024 Feb 29;15(1):1887. doi:10.1038/s41467-024-46142-w

25. Chen C, Chen B, Yang J, Li X, Peng X, Feng Y, et al. Development and validation of a practical machine learning model to predict sepsis after liver transplantation. Annals of Medicine. 2023;55(1):624–33. doi:10.1080/07853890.2023.2179104

26. Van De Sande D, Van Genderen ME, Smit JM, Huiskens J, Visser JJ, Veen RER, et al. Developing, implementing and governing artificial intelligence in medicine: a step-by-step approach to prevent an artificial intelligence winter. BMJ Health Care Inform. 2022 Feb;29(1):e100495. doi:10.1136/bmjhci-2021-100495

27. Cao C, Wu Y, Fang XZ, Liang Z, Mamykina L, Sbaffi L, et al. MedAI-SciTS: Enhancing Interdisciplinary Collaboration between AI Researchers and Medical Experts. In: Proceedings of the 2025 CHI Conference on Human Factors in Computing Systems [Internet]. Yokohama Japan: ACM; 2025 [cited 2026 Mar 16]. p. 1–24. Available from: https://dl.acm.org/doi/10.1145/3706598.3713926 doi:10.1145/3706598.3713926

28. Hassan M, Kushniruk A, Borycki E. Barriers to and Facilitators of Artificial Intelligence Adoption in Health Care: Scoping Review. JMIR Hum Factors. 2024 Aug 29;11:e48633. doi:10.2196/48633

29. Owoyemi A, Osuchukwu J, Salwei ME, Boyd A. Checklist Approach to Developing and Implementing AI in Clinical Settings: Instrument Development Study. JMIRx Med. 2025 Feb 20;6:e65565–e65565. doi:10.2196/65565

30. Giebel GD, Raszke P, Nowak H, Palmowski L, Adamzik M, Heinz P, et al. Improving AI-Based Clinical Decision Support Systems and Their Integration Into Care From the Perspective of Experts: Interview Study Among Different Stakeholders. JMIR Med Inform. 2025 Jul 7;13:e69688. doi:10.2196/69688

31. Hak F, Guimarães T, Santos M. Towards effective clinical decision support systems: A systematic review. PLoS One. 2022 Aug 15;17(8):e0272846. doi:10.1371/journal.pone.0272846 PubMed PMID: 35969526; PubMed Central PMCID: PMC9377614.

32. Tokgöz P, Hafner J, Dockweiler C. Faktoren für die Implementierung von KI-basierten Entscheidungsunterstützungssystemen zur Antibiotikavorhersage im Krankenhaus – eine qualitative Analyse aus der Perspektive von ärztlichem Personal. Gesundheitswesen. 2023 Jul 14;85(12):1220–8. doi:10.1055/a-2098-3108 PubMed PMID: 37451276; PubMed Central PMCID: PMC10713341.

33. Wang F, Beecy A. Implementing AI Models in Clinical Workflows: A Roadmap. BMJ Evid Based Med. 2025 Sep 22;30(5):285–7. doi:10.1136/bmjebm-2023-112727 PubMed PMID: 38914450; PubMed Central PMCID: PMC11666800.

34. Peek N, Capurro D, Rozova V, van der Veer SN. Bridging the Gap: Challenges and Strategies for the Implementation of Artificial Intelligence-based Clinical Decision Support Systems in Clinical Practice. Yearb Med Inform. 2025 Apr 8;33(1):103–14. doi:10.1055/s-0044-1800729 PubMed PMID: 40199296; PubMed Central PMCID: PMC12020628.

35. Sutton RT, Pincock D, Baumgart DC, Sadowski DC, Fedorak RN, Kroeker KI. An overview of clinical decision support systems: benefits, risks, and strategies for success. npj Digit Med. 2020 Feb 6;3(1):17. doi:10.1038/s41746-020-0221-y

36. Schmidt J, Schutte NM, Buttigieg S, Novillo-Ortiz D, Sutherland E, Anderson M, et al. Mapping the regulatory landscape for artificial intelligence in health within the European Union. npj Digit Med. 2024 Aug 27;7(1):229. doi:10.1038/s41746-024-01221-6

37. Meszaros J, Huys I, Ioannidis JPA. Challenges in applying the EU AI act research exemptions to contemporary AI research. npj Digit Med. 2026 Jan 31. doi:10.1038/s41746-025-02263-0

38. Meszaros J, Minari J, Huys I. The future regulation of artificial intelligence systems in healthcare services and medical research in the European Union. Front Genet. 2022 Oct 4;13. doi:10.3389/fgene.2022.927721

39. van Kolfschooten H, van Oirschot J. The EU Artificial Intelligence Act (2024): Implications for healthcare. Health Policy. 2024 Nov;149:105152. doi:10.1016/j.healthpol.2024.105152 PubMed PMID: 39244818.

40. Madara JL, Miyamoto S, Dowdy SC, Greene SM, Tarrant J, Margolis PA. Transforming Health Care — Shared Commitments for a Learning Health System. N Engl J Med. 2025 Jul 10;393(2):192–7. doi:10.1056/NEJMsb2507600

41. Steel PAD, Wardi G, Harrington RA, Longhurst CA. Learning health system strategies in the AI era. npj Health Syst. 2025 Jun 17;2(1):21. doi:10.1038/s44401-025-00029-0

42. Collins GS, Moons KGM, Dhiman P, Riley RD, Beam AL, Van Calster B, et al. TRIPOD+AI statement: updated guidance for reporting clinical prediction models that use regression or machine learning methods. BMJ. 2024 Apr 16;e078378. doi:10.1136/bmj-2023-078378

43. Jacob C, Brasier N, Laurenzi E, Heuss S, Mougiakakou SG, Cöltekin A, et al. AI for IMPACTS Framework for Evaluating the Long-Term Real-World Impacts of AI-Powered Clinician Tools: Systematic Review and Narrative Synthesis. J Med Internet Res. 2025 Feb 5;27:e67485. doi:10.2196/67485 PubMed PMID: 39909417; PubMed Central PMCID: PMC11840377.

44. van der Vegt AH, Scott IA, Dermawan K, Schnetler RJ, Kalke VR, Lane PJ. Implementation frameworks for end-to-end clinical AI: derivation of the SALIENT framework. J Am Med Inform Assoc. 2023 Sep 1;30(9):1503–15. doi:10.1093/jamia/ocad088

45. Nair M, Nygren J, Nilsen P, Gama F, Neher M, Larsson I, et al. Critical activities for successful implementation and adoption of AI in healthcare: towards a process framework for healthcare organizations. Front Digit Health. 2025 May 16;7. doi:10.3389/fdgth.2025.1550459

46. Reddy S, Rogers W, Makinen VP, Coiera E, Brown P, Wenzel M, et al. Evaluation framework to guide implementation of AI systems into healthcare settings. BMJ Health Care Inform. 2021 Oct 12;28(1):e100444. doi:10.1136/bmjhci-2021-100444 PubMed PMID: 34642177; PubMed Central PMCID: PMC8513218.

47. Lekadir K, Frangi AF, Porras AR, Glocker B, Cintas C, Langlotz CP, et al. FUTURE-AI: international consensus guideline for trustworthy and deployable artificial intelligence in healthcare. BMJ. 2025 Feb 5;388:e081554. doi:10.1136/bmj-2024-081554 PubMed PMID: 39909534.

48. Griesinger, Claudius Benedict, Reina, Vittorio, Panidis, Dimitrios, Chassaigne, Hubert. AI evidence pathway for operationalising trustworthy AI in health: an ontology unfolding ethical principles into translational and fundamental concepts [Internet]. Publications Office of the European Union; 2025 [cited 2026 May 20]. Available from: https://data.europa.eu/doi/10.2760/8107037

49. McLennan S, Fiske A, Tigard D, Müller R, Haddadin S, Buyx A. Embedded ethics: a proposal for integrating ethics into the development of medical AI. BMC Med Ethics. 2022 Dec;23(1):6. doi:10.1186/s12910-022-00746-3

50. Willem T, Fritzsche MC, Zimmermann BM, Sierawska A, Breuer S, Braun M, et al. Embedded Ethics in Practice: A Toolbox for Integrating the Analysis of Ethical and Social Issues into Healthcare AI Research. Sci Eng Ethics. 2024 Dec 24;31(1):3. doi:10.1007/s11948-024-00523-y

51. Ziethmann P, Hummel A, Althammer A, Schlögl-Flierl K, Heller AR, Brunner JO, et al. From Embedded Ethics to Explainable AI: Advancing Interdisciplinary Collaboration in Medical AI Development [Internet]. In Review; 2025 [cited 2025 Dec 28]. Available from: https://www.researchsquare.com/article/rs-7941605/v1 doi:10.21203/rs.3.rs-7941605/v1

52. Buschmeyer K, Hatfield S, Zenner J. Psychological assessment of AI-based decision support systems: tool development and expected benefits. Front Artif Intell. 2023 Sep 25;6. doi:10.3389/frai.2023.1249322

53. Gilbert S, Mathias R, Schönfelder A, Wekenborg M, Steinigen-Fuchs J, Dillenseger A, et al. A roadmap for safe, regulation-compliant Living Labs for AI and digital health development. Sci Adv. 2025 May 16;11(20):eadv7719. doi:10.1126/sciadv.adv7719

